# Psychological ownership influences perceived legal ownership of patient medical data

**DOI:** 10.1101/2023.04.03.23288075

**Authors:** Romain Cadario, Carmel Shachar, I. Glenn Cohen, Carey K. Morewedge

**Affiliations:** Erasmus University, Rotterdam School of Management; Harvard University, Harvard Law School; Boston University, Questrom School of Business

**Keywords:** **Keyworks**. Psychological ownership, legal ownership, medical decision making, data, privacy

## Abstract

We propose beliefs about legal ownership are informed and biased by intuitions about and cues that imbue psychological ownership—knowledge, control, and self-investment. We test our theory in the context of legal ownership of medical data in the United States, which is determined by property rights afforded by HIPAA to three stakeholders: patients, medical providers, and health systems. We theorize that stakeholders underestimate patient rights and overestimate provider rights afforded by HIPAA because patients are perceived to have less knowledge, control, and to have invested less in their data. As predicted, all stakeholders underestimated patient rights and overestimated provider rights afforded by HIPAA in a nationally representative sample of patients (*N*=300) and convenience samples of medical providers (*N*=114) and health systems administrators (*N*=100). All stakeholders agreed that patients should also be afforded more rights by HIPAA and fewer rights should be afforded to health systems. We find additional evidentiary support for our process account in three experiments (*N*=1195) in which we manipulated the perceived knowledge, control, and self-investment of patients, which modulated the degree to which patients underestimated rights afforded to them by HIPAA. Our findings illustrate how intuitions about psychological ownership inform and bias judgments of legal ownership, and reveal a consensus of stakeholders agree HIPAA should be reformed to expand patient rights.

**Public significance:** This study suggests that beliefs about legal ownership are informed and biased by intuitions about and cues that imbue psychological ownership—knowledge, control, and self-investment. It shows how patients, medical providers and health systems administrators underestimate patient rights and overestimate provider rights afforded by HIPAA, and reveals additional patient rights that all stakeholders agree should be afforded by HIPAA.

---

In the context of patient medical data, we test whether psychological ownership and the cues that imbue it inform and bias perceptions about legal ownership. Psychological ownership is the feeling that a thing is MINE. Legal ownership is generally attributed to persons or organizations who possess a “bundle” of property rights (Demsetz, 1974). We suggest that psychological ownership serves as an intuitive judgment of legal ownership when legal ownership is ambiguous (e.g., property rights are unknown). We test our theory with rights afforded by the Health Insurance Portability and Accountability Act (HIPAA) for patient medical data. Legal ownership of medical data includes property rights to use, control use, modify, sell, destroy, and transfer rights to other parties (Byrne, 2016; Oakley, 2023). HIPAA affords different bundles of property rights to three stakeholders: patients, medical providers, and health systems. The complexity of laws governing patient medical data often leaves stakeholders confused about these rights (Woelfel, 2005), which creates problems for data privacy, equity, and governance (Price & Cohen, 2019).

We suggest that psychological ownership informs and biases stakeholder beliefs about property rights for patient data. Three cues imbue psychological ownership from childhood to adulthood: perceived control, knowledge, and self-investment (Morewedge, 2021; Pierce et al., 2001). We theorize that the levels of these cues instantiate a bias in the perception of patient rights. Because relative to patients, medical providers are in a position of greater control, knowledge, and invest more resources into medical data than patients, rights afforded to medical providers are overestimated and rights afforded to patients are underestimated.

We first test our theory with a survey of US residents including a nationally representative sample of patients (N = 300) and convenience samples of medical providers (N = 114) and health systems administrators (N = 100). We find that stakeholders underestimate patient rights and overestimate medical provider rights afforded by HIPAA. This is not due to a confusion between perceived rights and perceived fairness. All stakeholders agree that HIPAA should afford additional rights to patients and reduce the rights afforded to health systems. In three experiments with US nationally representative samples, we manipulate the cues found to imbue psychological ownership—perceived knowledge, control, and self-investment (Morewedge, 2021; Pierce et al., 2001), and find these manipulations influence the degree to which patients underestimate the rights HIPAA affords them.

Our findings yield practical insights for the implementation, governance, and reform of HIPAA, health law and policy, and the regulation and sale of patient medical data. Our findings contribute to a literature examining effects of legal ownership on psychological ownership and downstream behaviors, such as the influence of stock ownership on psychological ownership and employee attitudes (Kim & Patel, 2024; Klein, 1987). We show that psychological ownership and the cues that imbue it resemble an intuitive ownership judgment that informs and biases judgments of legal ownership. Our findings also examine the influence of psychological ownership on legal ownership through a multiple stakeholder approach rather than from the perspective of individuals—whether a person feels an object or group is MINE or OURS (Jussila et al., 2015; Shu & Peck, 2011). We show how psychological ownership cues inform and bias perceptions of what is MINE and the property of others (i.e., “THEIRS”; Morewedge & Weiss, 2023).

## Psychological and Legal Ownership

Ownership can be legal, psychological, or both (Morewedge, 2021). Legal ownership is attributed to persons and organizations possessing one or more “sticks in a bundle” of property rights such as the property rights to use, control use, modify, sell, destroy, and transfer rights to other parties. Stakeholders can each be afforded or exchange rights. When “buying” a digital book, for instance, users purchase a long-term access right to the content. The platform and publishers and authors retain other rights. Psychological ownership is described as the feeling that a thing is MINE, which typically involves an association between possessions and the self (Belk, 1988; Ye & Gawronski, 2016). People feel psychological ownership for material, experiential, and abstract possessions (Dawkins et al., 2017; Pierce et al., 2001, 2003) from everyday household items to public parks and ideas (Morewedge & Giblin, 2015; Nancekivell et al., 2019; Ritov & Schurr, 2020). Psychological ownership often rises and falls with the acquisition and loss of legal ownership, such as when one acquires or sells a good or service (Marzilli Ericson & Fuster, 2014) and both are grounded in similar cues such as control and investment, but psychological and legal ownership are distinct concepts (Friedman et al., 2018). People feel psychological ownership for things they don’t legally own (e.g., companies in which they work) (Chen et al., 2023; Wang et al., 2019) and legally own things that they don’t feel are MINE (e.g., companies in which they hold stock).

We propose that psychological ownership and the cues that imbue it inform and bias perceptions of legal ownership, particularly when legal ownership is ambiguous. We test our theory by examining biases in perceived property rights afforded by HIPAA for patient medical data to three sets of stakeholders: patients, medical professionals, and health systems (e.g., hospitals and healthcare providers). Ownership of medical data is ambiguous and patients often do not know their legal rights (Balmer et al., 2010; Marshall & Barclay, 2003). As medical data is a product of patients who may know their health better than others (Belk, 1988), stakeholders may overestimate the property rights afforded to patients by HIPAA (Belk, 1988). We propose that healthcare providers should be more likely than patients to overestimate their rights, however, because providers experience higher levels of three cues to psychological ownership for patient data—perceived control (Nancekivell et al., 2019; Peck & Shu, 2009), subjective (medical) knowledge (Pierce et al., 2001, 2003), and self-investment (Kanngiesser et al., 2010; Norton et al., 2012).

Medical providers are typically the authority in patient-provider relationships (Popowicz, 2021), whereas patients generally lack control and autonomy (Goodyear-Smith & Buetow, 2001). Providers control patient medical data by collecting, classifying, reading, reporting, and modifying it. This dynamic is likely to bias perceived control and psychological ownership towards providers because, from early childhood, perceiving one has control over a thing creates a feeling that it is MINE (Espinosa & Starmans, 2020; Nancekivell et al., 2019). Control is also a critical factor in determining legal ownership, particularly in adverse possession cases where parties contest property rights (e.g., Pierson vs. Post, 1802). Feelings of control can be instantiated through direct physical contact, such as touching, holding, or repeatedly using an object, or by touching its virtual representation on a screen (Peck & Shu, 2009). Control can also be instantiated by determining who, when, and how others can access an object or experience (Neary et al., 2009). HIPAA allows patients to request a copy of their electronic health records, but few patients do, whereas providers regularly use patient electronic health records, for example, and patients may request changes to their electronic health records, but those changes are made by their providers. Second, medical providers are likely to possess greater professional knowledge and expertise than patients regarding the content and systems used to collect and maintain patient medical data. Greater subjective knowledge should imbue providers with a greater sense of psychological ownership over medical data than patients (Pierce et al., 2001, 2003). Third, patient medical data is generally contained in electronic health records. Providers and their employees typically invest more labor, time, and financial resources in the creation and maintenance of patient medical data than do patients. This kind of continued investment also has a parallel in adverse possession cases in the United States and Italy, where maintaining property can afford a person rights to that property.

We suggest that because psychological ownership serves as a form of intuitive legal ownership, the directionality of cues to psychological ownership inform and bias the rights that stakeholders believe are afforded by HIPAA to patients and medical providers. Given the higher perceived control, subjective knowledge, and self-investment experienced by medical providers than patients, we theorize that stakeholders exhibit biases consistent with these cues. We predict that stakeholders underestimate the rights afforded to patients and overestimate the rights afforded to medical providers by HIPAA. Furthermore, we predict that variation in psychological ownership cues—perceived control, subjective knowledge, and self-investment—modulate the severity of these biased beliefs.

## Transparency and Openness

We adhered to the *Journal of Experimental Psychology: General* methodological checklist. All manipulations and measures are reported for all studies. Study 1 was not pre-registered. The hypotheses, methods, and analysis plans were preregistered for Studies 2-4. The present research involved no more than minimal risks, and all study participants were 18 years of age or older. Informed consent was obtained for all participants. The study was approved for use with human participants by Institutional Review Boards of the first and last author. Pre-registrations, raw data, and R script files are available on our Open Science Framework repository at: https://osf.io/j7z26/?view_only=1aed05e5761b4a529efc270c5fa4c742.

## Study 1: Psychological Ownership Biases Legal Ownership

In Study 1, we tested our main predictions that stakeholders underestimate the number of rights afforded by HIPPA to patients and overestimate the number of rights afforded by HIPPA to medical providers. To inform health policy reform, we also examined biases in the perceived rights afforded by HIPPA to health systems.

## Methods

### Samples

We recruited a total of 514 American participants, divided into three samples. Through Survey Health Global we recruited 114 primary care physicians and 100 health systems administrators (e.g., hospitals). We recruited our patient sample through Lucid Theorem, a nationally representative sample of 300 Americans with respect to age, gender, ethnicity, and geography. Table 1 presents demographic variables for each sample, measured at the end of the survey (gender, age, health status, insurance provider, race, political orientation and income).

**Table 1.** Demographics by sample (Study 1)

| Variables | Patient sample<br>(N=300) | Medical provider<br>sample<br>(N = 114) | Health systems<br>administrators<br>sample<br>(N = 100) |
| --- | --- | --- | --- |
| Gender |  |  |  |
| Male | 47% | 67% | 52% |
| Female | 53% | 29% | 46% |
| Non-Binary | 0% | 2% | 0% |
| Prefer not to answer | 0% | 3% | 2% |
| Age |  |  |  |
| 18-24 | 12% | 0% | 1% |
| 25-34 | 22% | 4% | 2% |
| 35-44 | 17% | 18% | 25% |
| 45-54 | 17% | 27% | 34% |
| 55-64 | 16% | 30% | 28% |
| 65+ | 17% | 18% | 9% |
| Prefer not to answer | 0% | 4% | 1% |
| Income |  |  |  |
| <15k | 18% | 0% | 0% |
| 15k to 29.9k | 19% | 0% | 1% |
| 30k to 44.9k | 11% | 0% | 3% |
| 45k to 59.9k | 16% | 0% | 6% |
| 60k to 74.9k | 8% | 1% | 5% |
| 75k to 99.9k | 11% | 3% | 7% |
| 100k to 149.9k | 9% | 9% | 15% |
| 150k+ | 6% | 60% | 45% |
| Prefer not to answer | 3% | 29% | 18% |
| Race |  |  |  |
| Asian | 5% | 25% | 9% |
| Black | 10% | 4% | 3% |
| White | 74% | 57% | 79% |
| Other | 10% | 8% | 4% |
| Prefer not to answer | 1% | 6% | 5% |
| Insurance |  |  |  |
| Medicaid | 21% | 0% | 0% |
| Medicare | 31% | 12% | 7% |
| Private | 32% | 85% | 87% |
| Other | 7% | 2% | 4% |
| None | 8% | 1% | 0% |
| Prefer not to answer | 0% | 0% | 2% |
| Political orientation<br>(1: liberal to 7: conservative) | m = 4.43 | m = 4.61 | m = 4.83 |
| Health status<br>(1: poor to 5: excellent) | m = 3.12 | m = 4.03 | m = 4.07 |

### Measures

We measured perceptions of current and ideal property rights afforded by HIPAA to each group of stakeholders. We explicitly asked participants to evaluate the rights that each stakeholder *currently* possesses (i.e., perceived ownership) and *should* possess (i.e., ideal ownership) to make their distinction clear and address potential response bias. Measuring how property rights should ideally be afforded had the added benefit of identifying cases in which HIPAA and a stakeholder consensus diverge, informing health policy reform. Participants reported the current and ideal rights afforded by HIPAA to each of three stakeholder groups—patients, medical providers, and health systems in a 2 (ownership: perceived, ideal) x 6 (property rights) within-subject design. These questions are listed in Table 2 (for all stimuli see Supplementary Table 1). For each of the 12 questions, participants could select multiple stakeholders (e.g., “patients”, “medical providers” or “health systems”). Perceived and ideal rights afforded by HIPAA were presented in counterbalanced blocks.

**Table 2.** Perceived, ideal, and actual property rights afforded to stakeholders by HIPAA.

| Right # | Perceived ownership<br>Current condition [ideal condition] | Actual ownership |  |  |
| --- | --- | --- | --- | --- |
|  |  | Patients | Medical Providers | Health systems |
| Right #1:<br>Access | Subject to some exceptions, who *currently [should]* possesses the legal right to access a patient's electronic health records data (e.g., view it)? | ✓ | ✓ | ✓ |
| Right #2:<br>Control access | Subject to some exceptions, who *currently [should]* possesses the legal right to control access to a patient's electronic health record data (e.g., decide who can view it)? | ✓ | ✗ | ✓ |
| Right #3:<br>Profit | Subject to some exceptions, who *currently [should]* possesses the legal right to profit from or commercialize a patient's electronic health records data (e.g., by selling it)? | ✓ | ✗ | ✓ |
| Right #4:<br>Modify | Subject to some exceptions, who *currently [should]* possesses the legal right to modify a patient's electronic health records data (e.g., correct errors, make changes)? | ✓ | ✓ | ✓ |
| Right #5:<br>Destroy | Subject to some exceptions, who *currently [should]* possesses the legal right to destroy a patient's electronic health records data (e.g., delete all or part of it)? | ✗ | ✗ | ✗ |
| Right #6:<br>Transfer | Subject to some exceptions, who *currently [should]* possesses the legal right to transfer their property rights for a patient's electronic health records data (e.g., selling their own interest in that particular record)? | ✗ | ✗ | ✓ |
| Actual number of rights afforded to stakeholders by HIPAA |  | 4 | 2 | 5 |

Note: Actual ownership was determined by the second and third author, practicing lawyers with expertise in health policy law.

## Results

### Perceived vs. actual rights afforded by HIPAA

We first examined bias and accuracy by comparing (i) the perceived number of rights afforded to each group of stakeholders by HIPAA (white bars in Figure 1) and (ii) the actual number of rights legal afforded to each group of stakeholders by HIPAA (dashed line in Figure 1). The comparisons are plotted in Figure 1 for rights attributed to patients (Panel A), medical providers (Panel B), and health systems (Panel C). We used one-sample t-tests to identify departures from accuracy (i.e., 0); negative values indicate underestimation, and positive values indicate overestimation. We find a similar pattern of results when considering an alternative accuracy measure, the difference between the percentage of rights correctly afforded to each stakeholder and the actual percentage of rights afforded to each stakeholder (e.g., Supplementary Tables 2–4).

**Figure 1.**
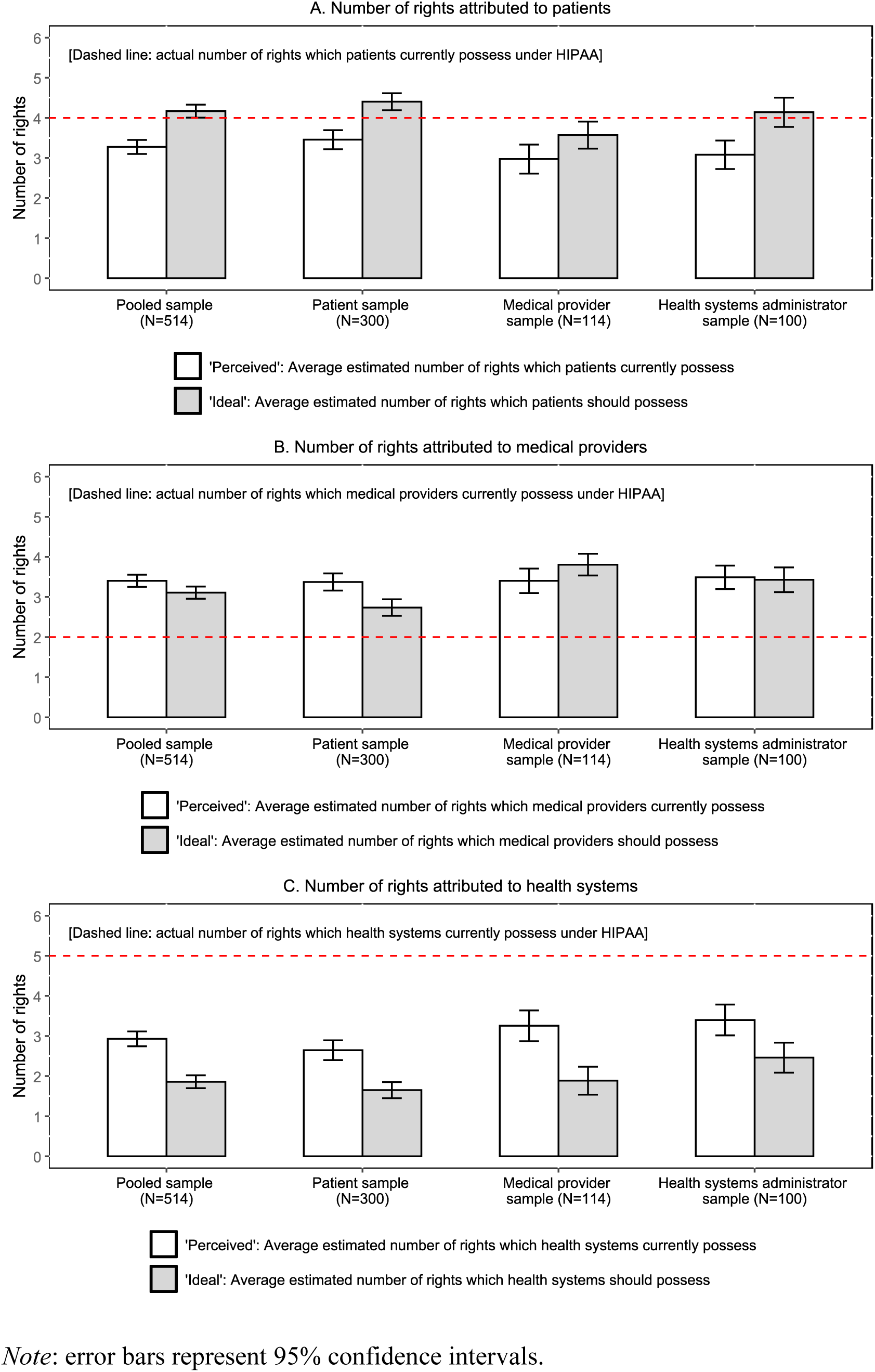
Perceived, ideal and actual legal rights by stakeholders (Study 1)

First, the pooled sample of all stakeholders (N=514) underestimated the rights that HIPAA affords to patients (*β* = −0.72, *t*(513) = −8.08, *p* < 0.001), overestimated the rights afforded by HIPAA to medical providers (*β* = 1.40, *t*(513) = 17.99, *p* < 0.001), and underestimated the rights afforded by HIPAA to health systems (*β* = −2.07, *t*(513) = −21.96, *p* < 0.001).

Second, patients (N=300) underestimated the rights that HIPAA affords them (*β* = - 0.53, *t*(299) = −4.46, *p* < 0.001). Patients overestimated the rights afforded by HIPAA to medical providers (*β* = 1.37, *t*(299) = 12.56, *p* < 0.001), and underestimated the rights afforded by HIPAA to health systems (*β* = −2.35, *t*(299) = −18.74, *p* < 0.001).

Third, medical providers (N=114) underestimated the rights that HIPAA affords to patients (*β* = −1.03, *t*(113) = −5.56, *p* < 0.001), overestimated the rights afforded by HIPAA to medical providers (*β* = 1.40, *t*(113) = 9.05, *p* < 0.001), and underestimated the rights afforded by HIPAA to health systems (*β* = −1.75, *t*(113) = −8.88, *p* < 0.001).

Fourth, health systems administrators (N=100) underestimated the rights that HIPAA affords to patients (*β* = −0.92, *t*(99) = −5.09, *p* < 0.001), overestimated the rights afforded by HIPAA to medical providers (*β* = 1.49, *t*(99) = 9.97, *p* < 0.001), and underestimated the rights afforded by HIPAA to health systems (*β* = −1.60, *t*(99) = −8.19, *p* < 0.001).

Last, we also report accuracy comparison between samples. Medical providers were more likely than patients to underestimate patient rights (*β* = −0.48, *t*(511) = −2.17, *p* = 0.030), but health systems administrators were no more likely than patients to underestimate patient rights (*β* = −0.38, *t*(511) = −1.61, *p* = 0.107). Providers and patients were similarly likely to overestimate medical provider rights (*p* = 0.877), as were health systems administrators and patients (*p* = 0.569). Underestimation of rights afforded to health systems was more pronounced in the provider than patient samples (*β* = 0.61, *t*(511) = 2.61, *p* = 0.009), and in the health systems administrator than patient samples (*β* = 0.75, *t*(511) = 3.08, *p* = 0.002).

### Perceived vs. ideal rights

As a comprehension check and to inform health policy reform, we compared the rights that participants believed HIPAA *does* afford (white bars in Figure 1) and the rights that participants believed HIPAA *should* afford to each stakeholder group by sample (gray bars in Figure 1). All stakeholders believed that patients should have more rights than they believe are currently afforded by HIPAA (*m_should_* = 4.17 vs. *m_current_* = 3.28, *t*(513) = 10.25, *p* < .001). There was no consensus about the rights medical providers should be afforded. Patients believed medical providers should have fewer rights than they believe providers are currently afforded (*m_should_* = 2.74 vs. *m_current_* = 3.37, *t*(299) = −5.25, *p* < .001). Medical providers believed they should have more rights than providers are currently afforded (*m_should_* = 3.81 vs. *m_current_* = 3.40, *t*(113) = 2.85, *p* = .005), and health systems administrators believed medical providers should have as many rights as they believed providers are currently afforded (*m_should_* = 3.43 vs. *m_current_* = 3.49, *t*(99) = −.39, *p* = .699). Paired sample t-tests revealed agreement among all stakeholders that health systems should have *fewer* rights than they are currently afforded (*m_should_* = 1.86 vs. *m_current_* = 2.93, *t*(99) = − 5.65, *p* < .001).

## Discussion

Patients, doctors, and administrators in Study 1 exhibited biases consistent with a psychological ownership framework in the rights they believe HIPAA affords to doctors and patients. Patients, doctors, and health systems administrators all overestimated the rights HIPAA affords to doctors, who are in a greater position of control, knowledge, and self-investment, and underestimated the rights that HIPAA affords to patients. Participants appeared to understand these assessments concerned the present rights of stakeholders rather than what they believe to be fair, as the perceived rights afforded to stakeholders differed from the ideal rights afforded to stakeholders.

### Studies 2-4: Cues to Psychological Ownership Influence Perceived Legal Ownership

In Studies 2-4, we focus on patient rights to directly test our process account. We experimentally manipulate cues that imbue psychological ownership (i.e., knowledge, control, and self-investment) and test the influence of these manipulations on the number of legal rights that patients believe are afforded to them by HIPAA.

### Study 2: Perceived Control and Legal Ownership

In Study 2, we examined how manipulating perceived control over electronic health records influences the perceived rights afforded to patients by HIPAA. We predicted that patients would believe fewer rights are afforded to them by HIPAA when patients felt less than more control over their electronic health records.

## Method

### Manipulation Pretest

We requested 200 and recruited 202 participants for a pretest from Prolific Academic (*m_age_* = 40.17, 64% female). In a between-subjects design, participants were randomly assigned to a high or low control condition in which they completed manipulations adapted from Whitson and Galinsky (2008). In the high control condition, we asked participants to, “*Write about the ways in which you have control in creating, accessing, and maintaining your electronic medical records.*” In the low control condition we asked participants to, “*Write about the ways your healthcare providers have control in creating, accessing, and maintaining your electronic medical records.*” All responses were recorded in an open-ended text box (see full stimuli in Supplementary Table 5). We measured perceived control over electronic health records on a 7-point scale with endpoints, 1 (not at all) and 7 (completely). Perceived control was greater in the high control condition than in the low control condition (*m_high control_* = 4.04, *se* = 0.18, *m_low control_* = 3.10, *se* = 0.16, *β* = −0.94, *t*(200) = −3.87, *p* < 0.001). See Table 3.

**Table 3.** Summary of the results from Studies 2-4.

|  | Study 2 | Study 3 | Study 4 |
| --- | --- | --- | --- |
|  | Control | Knowledge | Self-investment |
| Manipulation pretests: *N* | 202 | 200 | 199 |
| Low cue condition: *Mean* (*se*) | 3.10 (0.16) | 3.82 (0.11) | 2.74 (0.12) |
| High cue condition: *Mean* (*se*) | 4.04 (0.18) | 4.01 (0.10) | 3.21 (0.12) |
| Mean difference: *MD* (*p*-value) | 0.94 ($p < .001$) | 0.19 ($p = 0.011$) | 0.47 ($p = .006$) |
| Focal Experiments: *N* | 401 | 400 | 394 |
| Low cue condition: *Mean* (*se*) | 3.44 (0.12) | 3.50 (0.13) | 3.77 (0.11) |
| High cue condition: *Mean* (*se*) | 3.84 (0.13) | 3.87 (0.12) | 4.19 (0.12) |
| Mean difference<sup>a</sup>: *MD* (*p*-value) | 0.44 ($p = 0.020$) | 0.36 ($p = 0.039$) | 0.42 ($p = 0.012$) |

<sup>a</sup> *Pre-registered analysis.*

### Focal Experiment

We requested 400 participants and recruited 401 participants from a US nationally representative sample on Prolific Academic (*m_age_* = 45.78, 50% female). In a between-subjects design, participants were randomly assigned to a high or low control condition in which they completed the corresponding manipulations described in the pretest. All participants then reported their beliefs about the legal rights attributed to patients, medical providers, and health systems, as in Study 1.

## Results

As preregistered, we regressed the number of legal rights afforded to patients on the perceived control manipulation (1 = high control condition, 0 = low control condition). As predicted, participants believed HIPPA affords fewer rights to patients in the low control condition (*m_low control_* = 3.44, *se* = 0.12) than in the high control condition (*m_high control_* = 3.84, *se* = 0.13, *β* = −0.40, *t*(399) = −2.34, *p* = 0.020). Exploratory analyses reveal that participants also believed HIPPA affords more rights to medical providers in the low control condition (*m_low_* = 3.65, *se* = 0.11) than in the high control condition (*m_high_* = 3.34, *se* = 0.11, *β* = 0.31, *t*(399) = 2.04, *p* = 0.042).

### Study 3: Subjective Knowledge and Legal Ownership

In Study 3, we examined how manipulating subjective knowledge of electronic health records influences the perceived rights afforded to patients by HIPAA. We predicted that patients would believe fewer rights are afforded to them by HIPAA when patients felt less than more knowledgeable about their electronic health records.

## Method

### Manipulation Pretest

We requested and recruited 200 participants for a pretest from Prolific Academic (*m_age_* = 39.53, 56% female). We measured subjective knowledge of electronic health records (“How well do you feel like you understand how your electronic health records are created, stored, and managed?”) on a 7-point scale with endpoints, 1 (not well at all) and 7 (extremely well). Participants reported subjective knowledge before and after completing an intervention adapted from those used to reduce illusory subjective understanding (Cadario et al., 2021; Fernbach et al., 2013; Rozenblit & Keil, 2002).

> “We would like you to explain how your electronic health records are created, stored, and managed. Creation: Who creates your electronic health records? Storage: Where are your electronic health records stored? Management: Who can access and determine access to your electronic health records? Who can modify your electronic health records? Please describe all the details about each question.”

As expected, the manipulation reduced self-assessed subjective knowledge of electronic health records (*m_before_* = 4.01, *se_before_* = 0.10, *m_after_* = 3.82, *se_after_* = 0.11, *t*(199) = −2.56, *p* = 0.011).

### Focal Experiment

We requested and recruited 400 participants from a US nationally representative sample on Prolific Academic (*m_age_* = 45.76, 49% female). In a between-subjects design, participants were randomly assigned to a low or high perceived knowledge condition. In the low knowledge (treatment) condition, participants completed the manipulation designed to reduce subjective knowledge of electronic health records (see Supplementary Table 6). In the high knowledge condition, participants provided no explanation of their understanding of electronic health records; they immediately completed the dependent measures. All participants then reported their beliefs about the legal rights attributed to patients, medical providers, and health systems, as in Study 1.

## Results

As preregistered, we regressed the number of legal rights afforded to patients on the subjective knowledge manipulation (1 = low knowledge condition, 0 = high knowledge condition). As predicted, participants believed HIPPA affords fewer rights to patients in the low knowledge condition (*m_low knowledge_* = 3.50, *se* = 0.13) than in the high knowledge condition (*m_high knowledge_* = 3.87, *se*= 0.12, *β* = −0.36, *t*(397) = −2.07, *p* = 0.039). Exploratory analyses reveal that participants also believed HIPPA affords more rights to medical providers in the low knowledge condition (*m_low knowledge_* = 3.44, *se* = 0.11) than in the high knowledge condition (*m_high knowledge_* = 3.14, *se* = 0.10, *β* = 0.30, *t*(397) = 2.03, *p* = 0.043).

### Study 4: Self-Investment and Legal Ownership

In Study 4, we examined how manipulating perceived self-investment in electronic health records influences the perceived rights afforded to patients by HIPAA. We predicted that patients would believe fewer rights are afforded to them by HIPAA when patients felt they had invested less rather than more resources in their electronic health records.

## Method

### Manipulation Pretest

We requested 200 and recruited 199 participants for a pretest from Prolific Academic (*m_age_* = 35.34, 56% female). In a between-subjects design, participants were randomly assigned to a high or low self-investment condition. In the high self-investment condition, we asked participants to “*Write about the ways in which you have invested time and effort in creating, accessing, and maintaining your electronic medical records*” followed by a text box. In the low self-investment condition, we asked participants to “*Write about the ways your healthcare providers have invested time and effort in creating, accessing, and maintaining your electronic medical records*” (see full stimuli in Supplementary Table 7). We measured perceived investment by patients, providers, and health systems (“To what extent have each of the following groups below invested effort in creating, accessing, and maintaining your electronic medical records?”) on a 5-point scale with endpoints, 0 (none) and 4 (substantial effort). Perceived self-investment was lower in the low self-investment condition (*m_low_* = 2.74, *se_low_* = 0.12) than in the high self-investment condition (*m_high_* = 3.21, *se_high_* = 0.12, *β* = −0.47, *t*(197) = −2.79, *p* = 0.006).

### Focal Experiment

We requested 400 participants and recruited 394 participants from a US nationally representative sample on Prolific Academic (*m_age_* = 45.93, 50% female). In a between-subjects design, participants were randomly assigned to a low or high self-investment condition. In the low self-investment condition, participants were asked to “*Write about the ways your healthcare providers have invested time and effort in creating, accessing, and maintaining your electronic medical records*” (see full stimuli in Supplementary Table 7). In the high self-investment condition, participants were asked to “*Write about the ways in which you have invested time and effort in creating, accessing, and maintaining your electronic medical records.*” Open-ended responses were recorded in a text box. All participants then reported their beliefs about the legal rights attributed to patients, medical providers, and health systems, as in Study 1.

## Results

As preregistered, we regressed the number of legal rights afforded to patients on the self-investment manipulation (1 = high self-investment condition, 0 = low self-investment condition). As predicted, participants believed HIPAA affords fewer rights to patients in the low self-investment condition (*m_low self-investment_* = 3.77, *se_l_* = 0.11) than in the high self-investment condition (*m_high self-investment_* = 4.19, *se* = 0.12, *β* = −0.42, *t*(392) = −2.52, *p* = 0.012). Exploratory analyses revealed that participants did not believe HIPAA affords significantly more rights to medical providers in the low self-investment condition (*m_low self-investment_* = 3.33, *se* = 0.10) than in the high self-investment condition (*m_high self-investment_* = 3.11, *se* = 0.10, *β* = 0.22, *t*(392) = 1.60, *p* = 0.111), although those these means were directionally consistent with the predictions of our theory.

## Discussion

Cues giving rise to psychological ownership influenced perceived legal ownership of patient medical data. Manipulating perceived control, knowledge, and self-investment—cues that reliably induce psychological ownership (Morewedge, 2021; Pierce et al., 2001)— influenced the number of rights patients believed are afforded to them by HIPAA for their medical data. Manipulations that increased control, knowledge, and self-investment increased the number of rights patients believed are afforded to them by HIPAA, and manipulations that decreased these factors decreased the number of rights patients believed are afforded to them by HIPAA. As HIPAA affords four rights to patients, reducing perceived control, knowledge, and self-investment engendered more biased beliefs among patients.

### General Discussion

The rights stakeholders believe are afforded by HIPAA suggest that beliefs about legal ownership are informed and biased by intuitions about psychological ownership. As predicted by our psychological ownership framework, in a survey of 514 participants, including 300 patients, 114 providers, and 100 hospital administrators, stakeholders overestimated the number of rights afforded to medical providers by HIPPA and underestimated the number of rights afforded to patients. In three subsequent experiments, experimentally manipulating cues that imbue psychological ownership (i.e., knowledge, control and self-investment) affected the extent to which patients underestimated the rights afforded to them by HIPPA.

In unique samples, our findings demonstrate that cues to psychological ownership can inform and bias perceptions of legal ownership in contexts where legal ownership is consequential. In a literature that typically examines how legal ownership influences psychological ownership (Marzilli Ericson & Fuster, 2014; Morewedge & Giblin, 2015), our findings provide evidentiary support for a theory that psychological ownership serves as an intuitive judgment of legal ownership, which informs and biases judgments of legal ownership when it is ambiguous (Morewedge, 2021).

Cues to psychological ownership underlie the different perspectives of interdependent stakeholders. Research exploring connections between legal and psychological ownership typically examines perceptions from the viewpoint of a single stakeholder (Morewedge & Weiss, 2023). By investigating how multiple stakeholders (i.e., patients, medical providers, and health systems administrators) perceive ownership of medical data, we find evidence that different cue levels can create different perceptions of ownership across groups. One avenue for further research is whether rights afforded to stakeholders are assumed to be zero-sum, as our experiments found that manipulating cues to psychological ownership influenced the rights afforded to patients and doctors in opposing directions. Our approach highlights the complexities of ownership in multi-stakeholder environments and underscores the importance of considering multiple stakeholders and their relationships in psychological ownership research.

## Practical Implications

### Educational interventions to inform stakeholders about their rights

We find patients are unaware of HIPAA rights they are free to exercise. Our findings also suggest that medical providers and health systems administrators inaccurately inform patients about their rights. In Study 1, all three stakeholder groups overestimated the rights HIPAA affords to medical providers and underestimated the rights HIPAA affords to patients. All three groups also underestimated the rights HIPAA affords to health systems. All three stakeholder groups might benefit from educational interventions that better inform them about the rights that HIPAA affords to stakeholders, such as including a summary of patient rights in HIPAA authorization forms.

### Consensus among stakeholders for potential HIPAA reforms

So far, we considered the number of rights as our main dependent variable. In additional analyses from Study 1, we compared rights ideally afforded to each group of stakeholders by HIPAA and actual rights legal afforded to each group of stakeholders by HIPAA (Table 4).

**Table 4.** Perceived, ideal and actual legal rights attributed to stakeholders (Study 1)

|  | X = “Patients” |  |  | X = “Medical providers” |  |  | X = “Health systems” |  |  |
| --- | --- | --- | --- | --- | --- | --- | --- | --- | --- |
|  | Patient sample<br>(N=300) | Medical provider sample<br>(N=114) | Health system administrator<br>sample (N=100) | Patient sample<br>(N=300) | Medical provider sample<br>(N=114) | Health system administrator<br>sample (N=100) | Patient sample<br>(N=300) | Medical provider sample<br>(N=114) | Health system administrator<br>sample (N=100) |
| #1. Right to access a patient's EHR data | (✓) | (✓) | (✓) | (✓) | (✓) | (✓) | (✓) | (✓) | (✓) |
| X <u>currently</u> possess that right | 73% | 82% | 91% | 73% | 92% | 96% | 50% | 65% | 77% |
| X <u>should</u> possess that right | 83% | 82% | 89% | 65% | 97% | 93% | 42% | 51% | 69% |
| #2. Right to control access to a patient's EHR data | (✓) | (✓) | (✓) | (✗) | (✗) | (✗) | (✓) | (✓) | (✓) |
| X <u>currently</u> possess that right | 64% | 59% | 59% | 55% | 61% | 55% | 43% | 58% | 59% |
| X <u>should</u> possess that right | 79% | 65% | 77% | 41% | 75% | 53% | 21% | 36% | 41% |
| #3. Right to profit from patient's EHR data | (✓) | (✓) | (✓) | (✗) | (✗) | (✗) | (✓) | (✓) | (✓) |
| X <u>currently</u> possess that right | 47% | 43% | 35% | 43% | 22% | 24% | 49% | 59% | 59% |
| X <u>should</u> possess that right | 69% | 68% | 73% | 30% | 26% | 24% | 25% | 22% | 26% |
| #4. Right to modify a patient's EHR data | (✓) | (✓) | (✓) | (✓) | (✓) | (✓) | (✓) | (✓) | (✓) |
| X <u>currently</u> possess that right | 48% | 35% | 32% | 70% | 86% | 94% | 40% | 39% | 44% |
| X <u>should</u> possess that right | 60% | 31% | 44% | 66% | 93% | 92% | 28% | 20% | 43% |
| #5. Right to destroy a patient's EHR data | (✗) | (✗) | (✗) | (✗) | (✗) | (✗) | (✗) | (✗) | (✗) |
| X <u>currently</u> possess that right | 53% | 32% | 35% | 52% | 48% | 48% | 43% | 53% | 57% |
| X <u>should</u> possess that right | 70% | 50% | 57% | 38% | 52% | 49% | 28% | 32% | 40% |
| #6. Right to transfer rights for a patient's EHR data | (✗) | (✗) | (✗) | (✗) | (✗) | (✗) | (✓) | (✓) | (✓) |
| X <u>currently</u> possess that right | 60% | 47% | 56% | 44% | 32% | 32% | 40% | 53% | 44% |
| X <u>should</u> possess that right | 79% | 61% | 74% | 33% | 37% | 32% | 21% | 28% | 27% |

Note: (✓) means that the stakeholders X currently possess that right in reality, and (✗) means that the stakeholders X do not currently possess that right in reality. For each of the six legal rights, we report the percentage of participants who indicated that X (“patients”, “medical providers” or “healthy systems”) *currently possess* and *should possess* the right in question.

Overall, we find that all stakeholders agree that patients should be awarded two additional rights they are not presently afforded and that health systems should have fewer rights than they are presently afforded. Patients (70%), providers (50%), and administrators (57%) agree that patients should have the right to destroy their electronic health records (63% of the pooled sample). There is even greater agreement amongst patients (79%), providers (61%), and administrators (74%) that patients should have the right to transfer rights for their electronic health records (74% of the pooled sample). Interestingly, providers were the only sample who believe patients should lose a right they are presently afforded, with only 31% believing that patients should have the right to modify their electronic health records. For all comparisons, see Table 4 (left columns).

Levels of consensus were similarly high regarding the rights that should be taken away from health systems (Table 4, right columns). A minority of patients, providers, and administrators believed that health systems should have the right to control access to electronic health records (patients 21%, providers 36%, administrators 41%), to profit (patients 25%, providers 22%, administrators 26%), to modify patient electronic health records (patients 28%, providers 20%, administrators 43%), and the right to transfer rights (patients 21%, providers 28%, administrators 27%).

Less consensus was reached regarding rights afforded to medical providers by HIPAA (Table 4, middle columns). Providers believed they should be afforded four of six rights, including two rights they are presently afforded (to access 97% and modify 93% records) and two rights they are not afforded (control access 75%, destroy records 52%). Providers tentatively agreed (control access 53%, destroy 49%), but patients did not agree that providers should be afforded these additional rights (control access 41%, destroy records 38%).

In summary, our findings reveal a consensus among stakeholders for potential HIPAA reforms. All agree on expanding patient rights and curtailing the rights afforded to health systems. The US legal system typically supports data commercialization over privacy, making such policy changes challenging. The European Union’s General Data Protection Regulation (GDPR) is often considered a superior model for privacy (Forcier et al., 2019).

All stakeholders believed patients should have the right to destroy and transfer their medical data, rights not currently provided by HIPAA but afforded in some measure by the GDPR. This could involve adopting rights like the GDPR’s “right to be forgotten” and rethinking the monetization of patient data to better align with patients’ interests. Practically, it may be more difficult to curtail the rights of health systems but there does appear to be a consensus that those rights could be scaled back, even amongst health systems administrators.

### Framework for understanding biases in legal ownership

Finally, our findings suggest a framework for understanding biases in the inferences made by multiple stakeholders about the distribution of property rights. Our psychological ownership framework makes testable predictions by specifying the directional influence of cues including perceived control, familiarity and knowledge, and resource investment on the perception of property rights. Thus, our framework offers insights beyond health policy to more general inferences about property law and organizational governance.

## Limitations and Future Research

We note several limitations to consider when interpreting the findings. The sample size for medical providers and health system administrators was recruited through convenience sampling. This may limit the generalizability of the results to broader populations within these groups (e.g., medical providers can also include nurses, technicians, and specialists). Future research should aim to replicate these findings with larger and more diverse samples to ensure robustness and wider applicability.

Future research should delve deeper into the implications of psychological ownership for patient outcomes and healthcare practices. For example, understanding how overestimation of rights by providers and underestimation by patients affects trust, data sharing, and overall patient care could provide valuable insights for policymakers and healthcare professionals. Investigating interventions that align perceptions with actual legal rights might improve patient engagement, data security, and healthcare delivery.

## Conclusion

In conclusion, our studies reveal biases in how stakeholders perceive legal ownership of patient medical data. Medical providers overestimate the number of rights afforded to them under HIPAA while patients underestimate the number of rights afforded to them under HIPAA. Our findings highlight how psychological ownership, driven by cues such as control, knowledge, and self-investment, informs and biases believes about legal ownership. All stakeholders, including patients, providers, and administrators, agreed that patients should have more rights, suggesting potential for HIPAA reform. The results offer practical implications for health policy, emphasizing the need for educational interventions to clarify stakeholder rights.

## Author note

### Conflicts of interest

Prof. Cohen is chair of the ethics advisory board for Illumina and a member of the Bayer Bioethics Council. He was also compensated for speaking at events organized by Philips with the Washington Post and the Doctors Company, attending the Transformational Therapeutics Leadership Forum organized by Galen Atlantica, and retained as an expert in health privacy, gender-affirming care, and reproductive technology lawsuits. Funding: This research was supported by a grant from the Digital Business Institute in the Questrom School of Business at Boston University to Carey K. Morewedge. Pre-registrations, raw data and R script files are available on our Open Science Framework repository at: https://osf.io/j7z26/?view_only=1aed05e5761b4a529efc270c5fa4c742.

## Data Availability

https://osf.io/j7z26/?view_only=1aed05e5761b4a529efc270c5fa4c742.

## Supplementary Information

**Supplementary Table 1.**
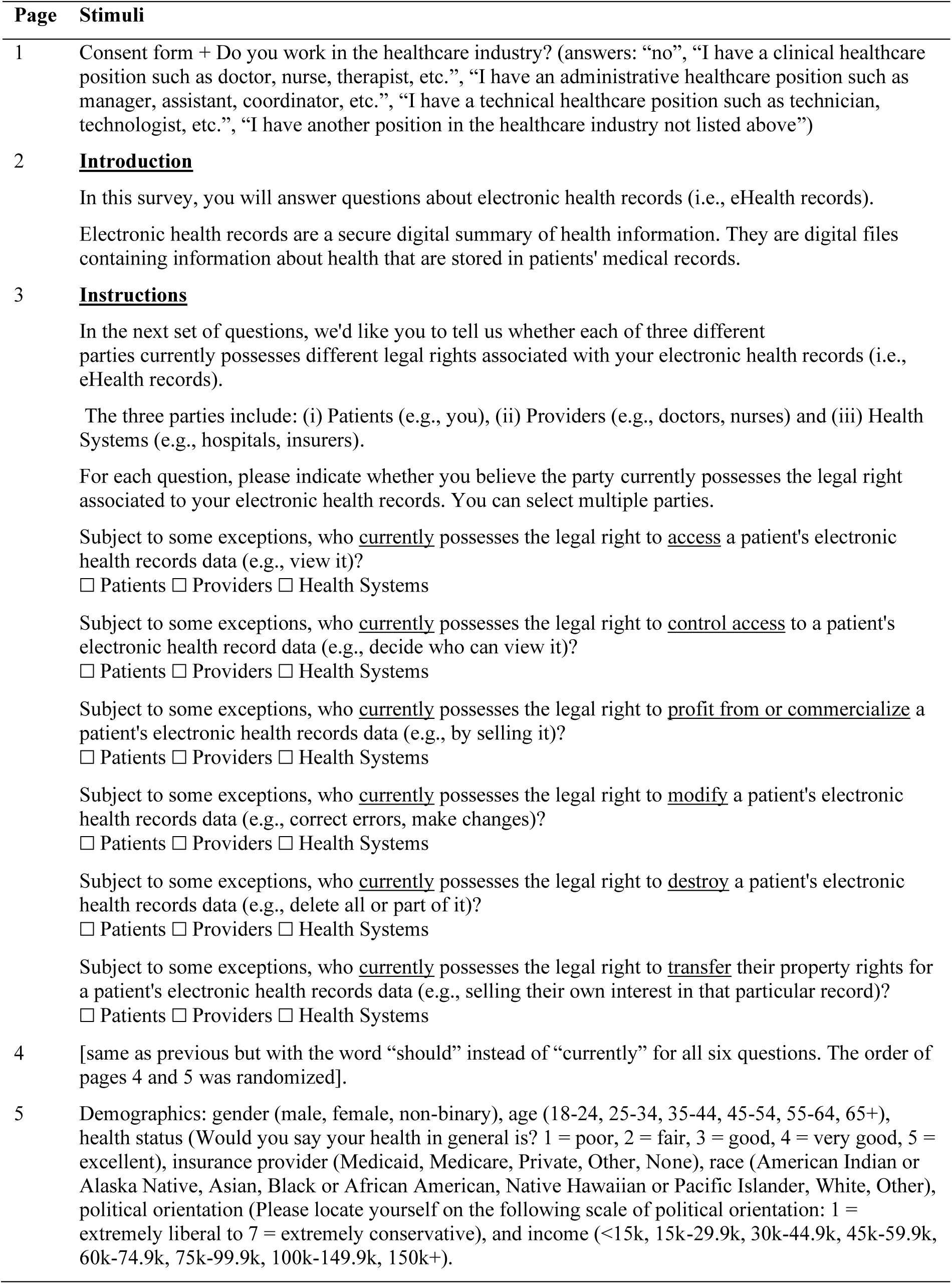
Stimuli for Study 1.

| Page | Stimuli |
| --- | --- |
| 1 | Consent form + Do you work in the healthcare industry? (answers: “no”, “I have a clinical healthcare position such as doctor, nurse, therapist, etc.”, “I have an administrative healthcare position such as manager, assistant, coordinator, etc.”, “I have a technical healthcare position such as technician, technologist, etc.”, “I have another position in the healthcare industry not listed above”) |
| 2 | **<u>Introduction</u>**<br>In this survey, you will answer questions about electronic health records (i.e., eHealth records).<br>Electronic health records are a secure digital summary of health information. They are digital files containing information about health that are stored in patients' medical records. |
| 3 | **<u>Instructions</u>**<br>In the next set of questions, we'd like you to tell us whether each of three different parties currently possesses different legal rights associated with your electronic health records (i.e., eHealth records).<br>The three parties include: (i) Patients (e.g., you), (ii) Providers (e.g., doctors, nurses) and (iii) Health Systems (e.g., hospitals, insurers).<br>For each question, please indicate whether you believe the party currently possesses the legal right associated to your electronic health records. You can select multiple parties.<br>Subject to some exceptions, who <u>currently</u> possesses the legal right to <u>access</u> a patient's electronic health records data (e.g., view it)?<br>☐ Patients ☐ Providers ☐ Health Systems<br>Subject to some exceptions, who <u>currently</u> possesses the legal right to <u>control access</u> to a patient's electronic health record data (e.g., decide who can view it)?<br>☐ Patients ☐ Providers ☐ Health Systems<br>Subject to some exceptions, who <u>currently</u> possesses the legal right to <u>profit from or commercialize</u> a patient's electronic health records data (e.g., by selling it)?<br>☐ Patients ☐ Providers ☐ Health Systems<br>Subject to some exceptions, who <u>currently</u> possesses the legal right to <u>modify</u> a patient's electronic health records data (e.g., correct errors, make changes)?<br>☐ Patients ☐ Providers ☐ Health Systems<br>Subject to some exceptions, who <u>currently</u> possesses the legal right to <u>destroy</u> a patient's electronic health records data (e.g., delete all or part of it)?<br>☐ Patients ☐ Providers ☐ Health Systems<br>Subject to some exceptions, who <u>currently</u> possesses the legal right to <u>transfer</u> their property rights for a patient's electronic health records data (e.g., selling their own interest in that particular record)?<br>☐ Patients ☐ Providers ☐ Health Systems |
| 4 | [same as previous but with the word “should” instead of “currently” for all six questions. The order of pages 4 and 5 was randomized]. |
| 5 | Demographics: gender (male, female, non-binary), age (18-24, 25-34, 35-44, 45-54, 55-64, 65+), health status (Would you say your health in general is? 1 = poor, 2 = fair, 3 = good, 4 = very good, 5 = excellent), insurance provider (Medicaid, Medicare, Private, Other, None), race (American Indian or Alaska Native, Asian, Black or African American, Native Hawaiian or Pacific Islander, White, Other), political orientation (Please locate yourself on the following scale of political orientation: 1 = extremely liberal to 7 = extremely conservative), and income (<15k, 15k-29.9k, 30k-44.9k, 45k-59.9k, 60k-74.9k, 75k-99.9k, 100k-149.9k, 150k+). |

**Supplementary Table 2.** Rights attributed to patients by sample (Study 1)

|  | Patient<br>sample<br>(N=300) | Medical<br>provider<br>sample<br>(N=114) | Health<br>systems<br>admin.<br>sample<br>(N=100) | Pooled<br>sample<br>(N=514) |
| --- | --- | --- | --- | --- |
| #1 Right to access a patient's EHR data (✓) |  |  |  |  |
| Patients <u>currently</u> possess that right | 73% | 82% | 91% | 78% |
| Patients <u>should</u> possess that right | 83% | 82% | 89% | 84% |
| #2 Right to control access to a patient's EHR data (✓) |  |  |  |  |
| Patients <u>currently</u> possess that right | 64% | 59% | 59% | 62% |
| Patients <u>should</u> possess that right | 79% | 65% | 77% | 75% |
| #3 Right to profit from patient's EHR data (✓) |  |  |  |  |
| Patients <u>currently</u> possess that right | 47% | 43% | 35% | 44% |
| Patients <u>should</u> possess that right | 69% | 68% | 73% | 70% |
| #4 Right to modify a patient's EHR data (✓) |  |  |  |  |
| Patients <u>currently</u> possess that right | 48% | 35% | 32% | 42% |
| Patients <u>should</u> possess that right | 60% | 31% | 44% | 50% |
| #5 Right to destroy a patient's EHR data (✗) |  |  |  |  |
| Patients <u>currently</u> possess that right | 53% | 32% | 35% | 45% |
| Patients <u>should</u> possess that right | 70% | 50% | 57% | 63% |
| #6 Right to transfer rights for a patient's EHR data (✗) |  |  |  |  |
| Patients <u>currently</u> possess that right | 60% | 47% | 56% | 57% |
| Patients <u>should</u> possess that right | 79% | 61% | 74% | 74% |
| A = % of rights correctly attributed to patients | 53% | 57% | 54% | 54% |
| B = actual % of rights attributed to patients (4/6) | 66.7% | 66.7% | 66.7% | 66.7% |
| A-B (* p < 0.05) | -13%* | -10%* | -12%* | -12%* |

Note: (✓) means that patients currently possess that right in reality, and (✗) means that patients do not currently possess that right in reality. For each of the six legal rights, we report the percentage of participants who indicated that “patients” *currently possess* and *should possess* the right in question.

**Supplementary Table 3.** Rights attributed to medical providers (Study 1)

|  | Patient<br>sample<br>(N=300) | Medical<br>provider<br>sample<br>(N=114) | Health<br>systems<br>admin.<br>sample<br>(N=100) | Pooled<br>sample<br>(N=514) |
| --- | --- | --- | --- | --- |
| #1 Right to access a patient's EHR data (✓) |  |  |  |  |
| Medical providers <u>currently</u> possess that right | 73% | 92% | 96% | 82% |
| Medical providers <u>should</u> possess that right | 65% | 97% | 93% | 78% |
| #2 Right to control access to a patient's EHR data (✗) |  |  |  |  |
| Medical providers <u>currently</u> possess that right | 55% | 61% | 55% | 56% |
| Medical providers <u>should</u> possess that right | 41% | 75% | 53% | 51% |
| #3 Right to profit from patient's EHR data (✗) |  |  |  |  |
| Medical providers <u>currently</u> possess that right | 43% | 22% | 24% | 34% |
| Medical providers <u>should</u> possess that right | 30% | 26% | 24% | 28% |
| #4 Right to modify a patient's EHR data (✓) |  |  |  |  |
| Medical providers <u>currently</u> possess that right | 70% | 86% | 94% | 78% |
| Medical providers <u>should</u> possess that right | 66% | 93% | 92% | 77% |
| #5 Right to destroy a patient's EHR data (✗) |  |  |  |  |
| Medical providers <u>currently</u> possess that right | 52% | 48% | 48% | 50% |
| Medical providers <u>should</u> possess that right | 38% | 52% | 49% | 43% |
| #6 Right to transfer rights for a patient's EHR data (✗) |  |  |  |  |
| Medical providers <u>currently</u> possess that right | 44% | 32% | 32% | 39% |
| Medical providers <u>should</u> possess that right | 33% | 37% | 32% | 34% |
| A = % of rights correctly attributed to providers | 58% | 69% | 72% | 63% |
| B = actual % of rights attributed to providers (2/6) | 33.3% | 33.3% | 33.3% | 33.3% |
| A-B (* p < 0.05) | +25%* | +36%* | +39%* | +30%* |

Note: (✓) means that medical providers currently possess that right in reality, and (✗) means that medical providers do not currently possess that right in reality. For each of the six legal rights, we report the percentage of participants who indicated that “medical providers” *currently possess* and *should possess* the right in question.

**Supplementary Table 4.** Rights attributed to health systems (Study 1)

|  | Patient<br>sample<br>(N=300) | Medical<br>provider<br>sample<br>(N=114) | Health<br>systems<br>admin.<br>sample<br>(N=100) | Pooled<br>sample<br>(N=514) |
| --- | --- | --- | --- | --- |
| #1 Right to access a patient's EHR data (✓) |  |  |  |  |
| Health systems <u>currently</u> possess that right | 50% | 65% | 77% | 59% |
| Health systems <u>should</u> possess that right | 42% | 51% | 69% | 49% |
| #2 Right to control access to a patient's EHR data (✓) |  |  |  |  |
| Health systems <u>currently</u> possess that right | 43% | 58% | 59% | 49% |
| Health systems <u>should</u> possess that right | 21% | 36% | 41% | 28% |
| #3 Right to profit from patient's EHR data (✓) |  |  |  |  |
| Health systems <u>currently</u> possess that right | 49% | 59% | 59% | 53% |
| Health systems <u>should</u> possess that right | 25% | 22% | 26% | 25% |
| #4 Right to modify a patient's EHR data (✓) |  |  |  |  |
| Health systems <u>currently</u> possess that right | 40% | 39% | 44% | 40% |
| Health systems <u>should</u> possess that right | 28% | 20% | 43% | 29% |
| #5 Right to destroy a patient's EHR data (✗) |  |  |  |  |
| Health systems <u>currently</u> possess that right | 43% | 53% | 57% | 48% |
| Health systems <u>should</u> possess that right | 28% | 32% | 40% | 31% |
| #6 Right to transfer rights for a patient's EHR data (✓) |  |  |  |  |
| Health systems <u>currently</u> possess that right | 40% | 53% | 44% | 43% |
| Health systems <u>should</u> possess that right | 21% | 28% | 27% | 24% |
| A = % of rights correctly attributed to health systems | 46% | 54% | 54% | 49% |
| B = actual % of rights attributed to health systems (5/6) | 83.3% | 83.3% | 83.3% | 83.3% |
| A-B (* p < 0.05) | -37%* | -29%* | -29%* | -34%* |

Note: (✓) means that health systems currently possess that right in reality, and (✗) means that health systems do not currently possess that right in reality. For each of the six legal rights, we report the percentage of participants who indicated that “health systems” *currently possess* and *should possess* the right in question.

**Supplementary Table 5.** Stimuli for Study 2.

| Page | Stimuli |
| --- | --- |
| 1 | [High control condition] Write about the ways in which **you have control** in creating, accessing, and maintaining your electronic medical records.<br>[Low control condition] Write about the ways **your healthcare providers have control** in creating, accessing, and maintaining your electronic medical records.<br>(50 characters minimum, validation available after 30 seconds) [text box] |
| 2 | **<u>Instructions</u>**<br>In the next set of questions, we'd like you to tell us whether each of three different parties currently possesses different legal rights associated with your electronic health records (i.e., eHealth records).<br>The three parties include<br>- Patients (e.g., you)<br>- Providers (e.g., doctors, nurses)<br>- Health Systems (e.g., hospitals, insurers)<br>For each question, please indicate whether you believe the party currently possesses the legal right associated to your electronic health records. You can select multiple parties.<br>Subject to some exceptions, who <u>currently</u> possesses the legal right to <u>access</u> a patient's electronic health records data (e.g., view it)?<br>☐ Patients ☐ Providers ☐ Health Systems<br>Subject to some exceptions, who <u>currently</u> possesses the legal right to <u>control access</u> to a patient's electronic health record data (e.g., decide who can view it)?<br>☐ Patients ☐ Providers ☐ Health Systems<br>Subject to some exceptions, who <u>currently</u> possesses the legal right to <u>profit from or commercialize</u> a patient's electronic health records data (e.g., by selling it)?<br>☐ Patients ☐ Providers ☐ Health Systems<br>Subject to some exceptions, who <u>currently</u> possesses the legal right to <u>modify</u> a patient's electronic health records data (e.g., correct errors, make changes)?<br>☐ Patients ☐ Providers ☐ Health Systems<br>Subject to some exceptions, who <u>currently</u> possesses the legal right to <u>destroy</u> a patient's electronic health records data (e.g., delete all or part of it)?<br>☐ Patients ☐ Providers ☐ Health Systems<br>Subject to some exceptions, who <u>currently</u> possesses the legal right to <u>transfer</u> their property rights for a patient's electronic health records data (e.g., selling their own interest in that particular record)?<br>☐ Patients ☐ Providers ☐ Health Systems |
| 3 | Gender (male, female, non-binary), age (in years) |

**Supplementary Table 5.**
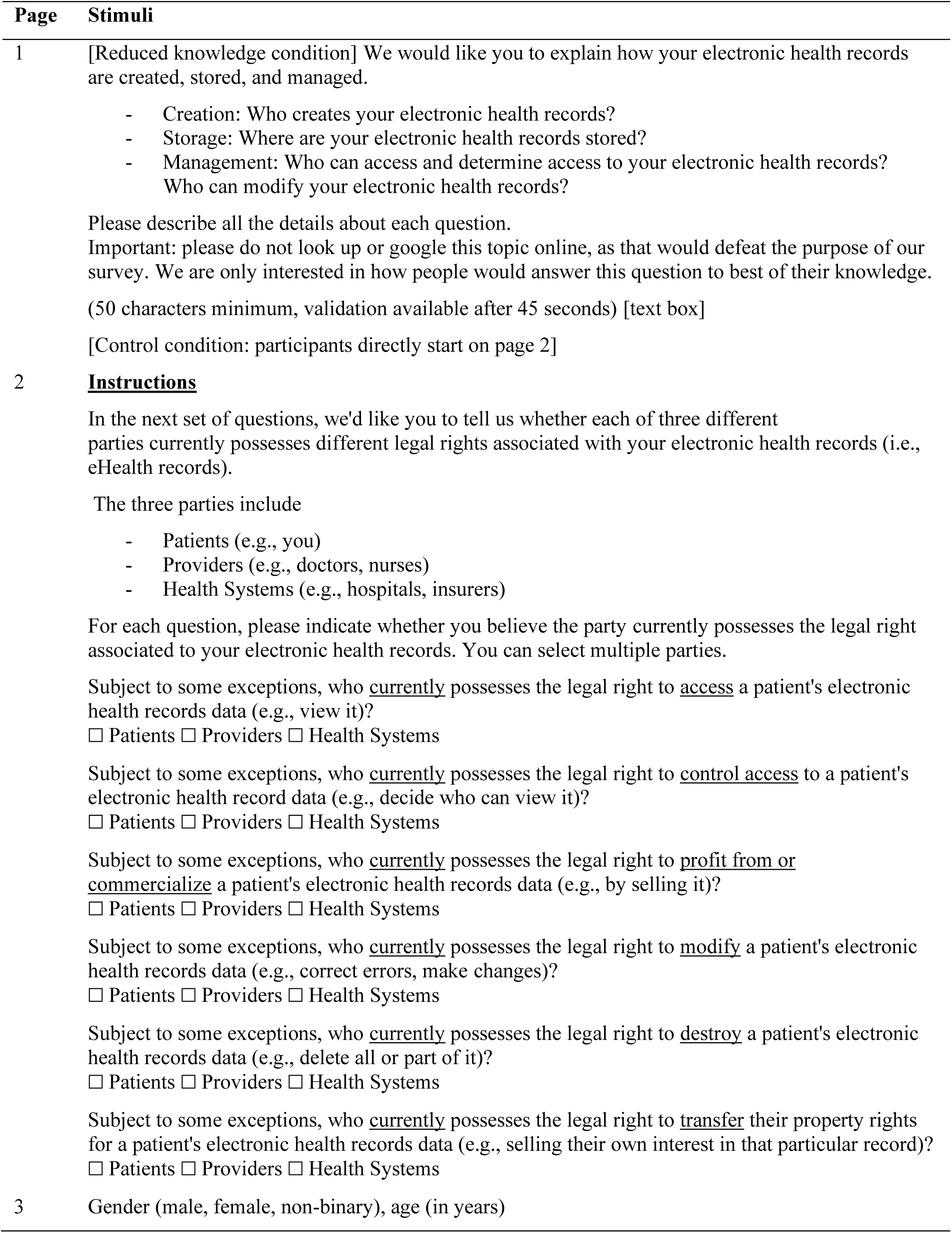
Stimuli for Study 3.

| Page | Stimuli |
| --- | --- |
| 1 | [Reduced knowledge condition] We would like you to explain how your electronic health records are created, stored, and managed.<br>- Creation: Who creates your electronic health records?<br>- Storage: Where are your electronic health records stored?<br>- Management: Who can access and determine access to your electronic health records? Who can modify your electronic health records?<br>Please describe all the details about each question.<br>Important: please do not look up or google this topic online, as that would defeat the purpose of our survey. We are only interested in how people would answer this question to best of their knowledge.<br>(50 characters minimum, validation available after 45 seconds) [text box]<br>[Control condition: participants directly start on page 2] |
| 2 | **<u>Instructions</u>**<br>In the next set of questions, we'd like you to tell us whether each of three different parties currently possesses different legal rights associated with your electronic health records (i.e., eHealth records).<br>The three parties include<br>- Patients (e.g., you)<br>- Providers (e.g., doctors, nurses)<br>- Health Systems (e.g., hospitals, insurers)<br>For each question, please indicate whether you believe the party currently possesses the legal right associated to your electronic health records. You can select multiple parties.<br>Subject to some exceptions, who <u>currently</u> possesses the legal right to <u>access</u> a patient's electronic health records data (e.g., view it)?<br>☐ Patients ☐ Providers ☐ Health Systems<br>Subject to some exceptions, who <u>currently</u> possesses the legal right to <u>control access</u> to a patient's electronic health record data (e.g., decide who can view it)?<br>☐ Patients ☐ Providers ☐ Health Systems<br>Subject to some exceptions, who <u>currently</u> possesses the legal right to <u>profit from or commercialize</u> a patient's electronic health records data (e.g., by selling it)?<br>☐ Patients ☐ Providers ☐ Health Systems<br>Subject to some exceptions, who <u>currently</u> possesses the legal right to <u>modify</u> a patient's electronic health records data (e.g., correct errors, make changes)?<br>☐ Patients ☐ Providers ☐ Health Systems<br>Subject to some exceptions, who <u>currently</u> possesses the legal right to <u>destroy</u> a patient's electronic health records data (e.g., delete all or part of it)?<br>☐ Patients ☐ Providers ☐ Health Systems<br>Subject to some exceptions, who <u>currently</u> possesses the legal right to <u>transfer</u> their property rights for a patient's electronic health records data (e.g., selling their own interest in that particular record)?<br>☐ Patients ☐ Providers ☐ Health Systems |
| 3 | Gender (male, female, non-binary), age (in years) |

**Supplementary Table 7.** Stimuli for Study 4.

| Page | Stimuli |
| --- | --- |
| 1 | [High self-investment condition] Write about the ways in which **<u>you</u>** have invested time and effort in creating, accessing, and maintaining your electronic medical records.<br>[Low self-investment condition] Write about the ways your **<u>healthcare providers</u>** have invested time and effort in creating, accessing, and maintaining your electronic medical records.<br>(50 characters minimum, validation available after 30 seconds) [text box] |
| 2 | **<u>Instructions</u>**<br>In the next set of questions, we'd like you to tell us whether each of three different parties currently possesses different legal rights associated with your electronic health records (i.e., eHealth records).<br>The three parties include<br>- Patients (e.g., you)<br>- Providers (e.g., doctors, nurses)<br>- Health Systems (e.g., hospitals, insurers)<br>For each question, please indicate whether you believe the party currently possesses the legal right associated to your electronic health records. You can select multiple parties.<br>Subject to some exceptions, who **<u>currently</u>** possesses the legal right to **<u>access</u>** a patient's electronic health records data (e.g., view it)?<br>☐ Patients ☐ Providers ☐ Health Systems<br>Subject to some exceptions, who **<u>currently</u>** possesses the legal right to **<u>control access</u>** to a patient's electronic health record data (e.g., decide who can view it)?<br>☐ Patients ☐ Providers ☐ Health Systems<br>Subject to some exceptions, who **<u>currently</u>** possesses the legal right to **<u>profit from or commercialize</u>** a patient's electronic health records data (e.g., by selling it)?<br>☐ Patients ☐ Providers ☐ Health Systems<br>Subject to some exceptions, who **<u>currently</u>** possesses the legal right to **<u>modify</u>** a patient's electronic health records data (e.g., correct errors, make changes)?<br>☐ Patients ☐ Providers ☐ Health Systems<br>Subject to some exceptions, who **<u>currently</u>** possesses the legal right to **<u>destroy</u>** a patient's electronic health records data (e.g., delete all or part of it)?<br>☐ Patients ☐ Providers ☐ Health Systems<br>Subject to some exceptions, who **<u>currently</u>** possesses the legal right to **<u>transfer</u>** their property rights for a patient's electronic health records data (e.g., selling their own interest in that particular record)?<br>☐ Patients ☐ Providers ☐ Health Systems |
| 3 | Gender (male, female, non-binary), age (in years) |

## Notes

### Clinical Protocols

https://osf.io/j7z26/?view_only=1aed05e5761b4a529efc270c5fa4c742.

### Author Declarations

The study was approved for use with human participants by the Institutional Review Boards of the Charles River Campus at Boston University (protocol 3632E) and of the Erasmus University Erasmus Research Institute of Management (ETH2122-0846).

### Summary of Updates

Revision to Study 1 and Figure 1.

